# A stepped-care programme of brief psychological interventions for adults affected by adversity in Jordan: Lessons from a pilot randomised controlled trial in Jordan

**DOI:** 10.1101/2025.02.27.25323060

**Authors:** Dharani Keyan, Rand Habashneh, Hafsa El-Dardery, Muhannad Faroun, Feda’a Al-Johary, Adnan Abualhaija, Ibrahim Said Aqel, Aemal Akhtar, Latefa Dardas, Richard A. Bryant

**Author notes:** Corresponding author: Dharani Keyan, PhD, School of Psychology, University of New South Wales, Sydney, NSW 2052 Australia.

## Abstract

**Background:** Stepped care frameworks comprise of the use of relatively limited resources to serve the greatest number of people.

**Objective:** We conducted a pilot randomised controlled trial in Jordan between September 2022 and February 2023 to evaluate the feasibility of a stepped care model of psychological care for distressed adults.

**Methods:** Participants were randomised to receive a stepped care intervention involving a guided self-help program (*Doing What Matters;* DWM), and if still met criteria for psychological distress, followed by a more intensive group program (group *Problem Management Plus*; gPM+) or a guided self-help program alone. Both intervention arms also received enhanced treatment as usual (ETAU) that comprised referral to available community support services. A mixed methods design was used to assess feasibility and acceptability and expected clinical outcomes.

**Findings:** One hundred and forty-five distressed adults (meeting Kessler Distress Score-10 score of ≥20) were randomised to either the stepped care or the single intervention arm (86% female) on a 1:1 allocation basis. DWM was delivered over five weeks by trained non-specialist helpers, where participants attended on average 2.6 support calls. Those randomised to gPM+ attended on average 3.3 sessions. The study demonstrated feasibility and acceptability for DWM and gPM+ interventions as delivered by trained non-specialists. Although the trial was not powered to detect clinical effectiveness, the stepped care arm relative to the single intervention arm demonstrated significantly lower depression symptoms immediately after and at 3-months following intervention delivery.

**Conclusions:** The study and trial procedures were acceptable to participants, non-specialists, and programme staff and demonstrated feasibility of implementing such a framework in Jordan.

**Clinical implications:** These findings informed a fully powered definitive RCT seeking to evaluate the clinical and cost-effectiveness of a stepped model of care in Jordan.

**What is already known on this topic:**

- Stepped care frameworks, involving the use of relatively limited resources to serve the greatest number of people, is one avenue to address the growing burden of common mental disorders in low- and middle-income countries (LMICs).

**What this study adds:**

- This is the first feasibility trial to demonstrate acceptability for a stepped model of scalable psychological care as delivered by trained non-specialists for distressed adults in Jordan.
- This is also the first trial to explore the feasibility and acceptability of the World Health Organization interventions involving the guided self-help intervention (*Doing What Matters*; DWM) followed *Problem Management Plus* (PM+) within a stepped care framework in an exemplar LMIC setting.

**How this study might affect research, practice or policy:**

- These findings informed a fully powered trial seeking to evaluate the clinical and cost-effectiveness of delivering brief, scalable psychological interventions in Jordan.

---

Attempts to reduce the burden of common mental disorders in low-middle-income countries (LMICs) come with the challenge of focusing on helping the greatest number of people with relatively limited resources. To overcome such resource barriers to mental health care, the World Health Organization (WHO) has developed affordable, brief, and non-specialist delivered interventions for communities affected by adversity. *Doing What Matters in Times of Stress* (DWM) is an adaptation of WHO’s evidence-based stress-management program, *Self-Help Plus* (SH+) ^1^, an early intervention that has been shown to be effective in preventing or reducing psychological distress ^2^ ^3^. DWM involves self-guided exercises in stress management that promotes one’s psychological flexibility based on techniques from acceptance and commitment therapy. These exercises are outlined in an illustrated booklet and are supported with 15-minute phone calls over five weeks ^4^. In comparison, the WHO’s *Problem Management Plus* (PM+) is a lay-facilitator led program that instructs techniques in slow breathing, behavioural activation, problem management and social support enhancement based on cognitive behavioural- and problem-solving therapies ^5^. Like DWM, PM+ is also delivered across a 5-week period in either individual or group format (gPM+), with reductions in psychological distress evident in diverse countries in both the individual ^6^ ^7^ and group ^8–10^ formats. Further, a recent meta-analysis has indicated that PM+ achieves a moderate effect on reducing psychological distress ^11^.

Despite the success of these programs, from the perspective of health system providers there is a need to ‘do more with less’. To this end, there is recognition that stepped care frameworks can be useful to provide very low intensity interventions to people who can benefit from them and restrict more labour-intensive programs to those with more persistent psychological needs. In the context of the WHO interventions, an initial step would be to provide access to DWM. Following this, individuals who do not report sufficient reductions in psychological distress (based on a pre-determined target milestone) would be stepped up to receive a relatively more intensive intervention, PM+, to address residual symptoms and problems. Although stepped care frameworks have been evidenced to be cost-effective and increasingly favored in high-income settings ^12^, there is a dearth of evidence for the efficacy of stepped care models in LMICs. There is a need to compare the relative efficacies of a stepped care program relative to a single self-guided intervention for brief psychological interventions in LMIC settings. To understand, the need, acceptability, and feasibility for such a framework in Jordan, an LMIC setting, we conducted a pilot randomized controlled trial (RCT) to evaluate a stepped care model of psychological interventions for distressed adults residing in the country. The aims of this study were to a) test trial procedures in preparation for a definitive RCT; b) evaluate acceptability and feasibility; and c) examine likely clinical effectiveness of this stepped model of care among Jordanians and refugees with elevated levels of psychological distress. These aims were outlined to inform the conduct of a subsequent fully powered effectiveness trial of a stepped care model of scalable psychological care in Jordan.

## Methods

### Setting

This study was carried out by the University of New South Wales (UNSW), Sydney and implemented by the Institute for Family Health (IFH) in Jordan. Jordan is a small middle-income country in the Middle East that has been heavily impacted by ongoing conflicts in the neighboring Arab regions over the past decade. Its social and health climate is increasingly characterized by high proportions of refugees, rising unemployment, poverty, and low access to health services including mental health ^13^. IFH is a national non-governmental organisation providing comprehensive health care including physical, mental, and social services in urban and regional areas for Jordanians and refugees residing across the country.

### Design

The pilot study was designed as a two-arm single blind randomized controlled trial (RCT), comparing a single intervention (i.e., DWM) v. a stepped care (i.e., DWM followed by gPM+). Details of the a priori aims and procedures have been detailed elsewhere ^14^.

### Recruitment and Participants

Adults (18 years or above) residing across two local governorate sites including Amman and Karak who were existing beneficiaries of IFH services were first contacted by a phone call to enquire about potential interest in participating in this pilot study. Those expressing interest provided verbal consent during this time and completed a screening measure (i.e., K10 assessment). Inclusion criteria were a) individuals with elevated psychological distress (as indicated by a score ≥ 20 on the Kessler Psychological Distress Scale (K10) ^15^ and b) reporting sufficient proficiency in the Arabic language. Individuals were excluded if they reported (a) imminent plans of suicide or risk to their personal safety (e.g., partner violence), (b) indicated severe mental disorders (i.e., psychosis), (c) had cognitive impairment, (d) plans to return to Syria in the next 12 months, or (e) had no access to a telephone.

### Randomization

Those meeting inclusion criteria were randomized using computer-generated sequences that were stratified based on demographic (ratio of 6:4) for Jordanian and refugee (including Syrian, Palestinian, Iraqi or other refugees) participants within each recruitment site (i.e., Amman and Karak) on a 1:1 basis. Participants were randomized to either stepped care (i.e., DWM + gPM*+/*Enhanced Treatment as Usual (ETAU)) or self-guided intervention alone (i.e., DWM/ETAU).

### Assessment procedures

Within eight working days of screening, participants were contacted by independent assessors to complete baseline assessments (T0). Within one week of T0 assessments, participants were randomized, contacted by their helper who later visited their homes to obtain written consent and provided them with the DWM booklet. Interim assessments (T1) were conducted immediately after completing the DWM intervention during which time, the local project coordinator communicated group allocation to participants. Post (T2) and 3- month follow-up (T3) assessments were scheduled one week-(six weeks since baseline), and 12 weeks after the 5^th^ PM+ session (18 weeks since baseline), respectively. All assessments were completed by assessors who were blind to randomization and situated in a location separate to helpers to maintain independence of assessors. Additionally, assessors indicated intervention arm assignment following each assessment stage (T0-T3) to verify whether the blind was broken. These assessors received a 4-day training consisting of administration of questionnaires, general interview techniques, and ethical research conduct. Participants were reimbursed 3 JOD upon completion of all assessments.

### Interventions

*Doing What Matters* (DWM): The self-guided intervention, DWM ^16^, consists of an illustrated booklet of five stress management strategies (including audio exercises) that participants work through in a self-paced manner over a period of five weeks. To support learning and practice of strategies, participants opted to receive three 15-minute phone calls at pre-specified time points (i.e., weeks 1, 3, & 5) during which time they received support on use of strategies and were guided with trouble shooting their practice. The adaptation of DWM for this study has been detailed elsewhere ^14^. DWM was translated into conversational Arabic prior to administration. Cultural adaption of content was completed by conducting focus groups with representative participants and clinical service providers at IFH to obtain their feedback on audio, visual, and written content in line with a standardized framework ^17^.

*Enhanced Treatment as Usual* (ETAU): Participants were provided with a single follow-up referral session over a phone call consisting of information on available support services within the community.

*Group Problem Management Plus* (gPM+): The facilitator led group intervention, gPM*+,* consisted of five weekly 90-minute sessions in groups of 8-10 individuals. This consisted of four strategies including a slow diaphragmatic breathing exercise, structured problem solving, behavioral activation across pleasurable and task-oriented activities and increasing access to social supports. A detailed description of these PM+ strategies is available elsewhere ^5^. Cultural adaptation of gPM+ has been previously conducted in this setting ^18^. Those participants residing at a distance from the IFH centers were reimbursed 3 JOD for their travel to attend gPM+ sessions.

### Main outcomes

The main outcomes that were the focus of this study were feasibility and acceptability including fidelity to- and drop-out from interventions and likely clinical effectiveness of the interventions within a stepped care framework.

### Qualitative evaluation

Evaluation of all study activities followed a semi-structured interview guide. Interviews were conducted with program staff and participants who had different rates of retention in the DWM and gPM+ interventions. Key informants included assessors, participants who completed and dropped out of the DWM and gPM+ the interventions, DWM helpers, gPM+ facilitators, and a clinical supervisor of both interventions. Interviews took place in both focus group discussions and individual key informant formats where questions explored the appropriateness and acceptability of interventions within the stepped care approach, challenges that were experienced during participation and suggestions to improve trial procedures.

### Quantitative measures

The primary clinical outcome was the Hopkins Symptom Checklist-25 (HSCL-25; ^19^) which measured anxiety and depression on a 4-point scale (where higher scores indicate increased severity). It has been clinically validated in cross-cultural settings including Jordan^20^. The WHO Well-Being Index (WHO-5; ^21^) was administered to assess states of positive mood, interest and energy.

Sociodemographic information was collected from questions A1-A5 of the WHO Disability Assessment Schedule 2.0 (WHODAS 2.0; ^22^). Exposure to potentially traumatic events was assessed using the Life Events Checklist (LEC; ^23^), a 17-item list of experienced or witnessed traumatic events (e.g., rape, serious injury, combat exposure or the sudden death of a loved one). The LEC was adapted to consist of a list 27 items based on qualitative assessments conducted in prior trials within LMIC settings ^8^.

Other secondary outcomes included the WHODAS 2.0 and EURO-QoL to obtain information relating to functioning and quality of life. The WHODAS 2.0 assesses difficulties experienced (because of illness or mental health conditions) across six domains of functioning including, cognition, mobility, self-care, getting along, life activities and participation during the past 30 days. The 12-item interviewer version scores a participant’s difficulties on a 5-point Likert scale, where higher scores indicate worse functional impairment. The EURO-QoL-5D-5L ^24^ assesses participants’ health across five domains (i.e., mobility, self-care, usual activities, pain/discomfort and anxiety/ depression) and five levels of functioning (no problems, slight problems, moderate problems, severe problems and unable to/extreme problems). Participants rate their overall health using a visual analogue scale.

Other secondary outcomes included the Psychological Outcomes Profiles (PSYCHLOPS; ^25^) to assess the main problems faced by participants. Participants are asked to self-identify problems prior-to the start of interventions, and later where they are asked to re-rate their endorsement of these problems at the end of the interventions. Other outcomes included self-reported agency as measured using the agency subscale of the State Hope Scale^26^, where this measure has been validated following brief behaviour change interventions ^27^.

### Quantitative and Qualitative analyses

Qualitative data from semi-structured interviews were audio recorded, transcribed verbatim, and then translated into English by a professional translator. The interviews were conducted by the project coordinator involved in study procedures. A combination of inductive (codes derived from the data) and deductive (codes derived from constructs related to feasibility) thematic analyses were used. All data were coded in NVivo version 14 ^28^. Two coders first familiarized themselves with the transcripts, and coded interviews based on inductive and deductive codes. Findings were discussed between coders and final coding framework was applied to all transcripts.

Baseline socio-demographic characteristics were descriptively summarized and analyzed using independent samples t-tests and chi-square tests. Although this study was not powered to detect statistically significant differences, we conducted linear mixed models to explore potential differential trajectories between conditions on primary and secondary clinical outcomes. Additionally, effect sizes for within intervention arms were calculated divided the mean differences between assessments (T0 to T3) by the pooled standard deviation (Cohen’s *d*). This was done to help inform a future fully powered RCT.

### Ethics

This study received local ethical approval from the University of Jordan (number: PF.22.10). This study was prospectively registered on 22 February 2021 (ACTRN12621000189820p). Participants were enrolled in the study after obtaining written consent.

## Results

### A) Testing trial procedures

#### Recruitment and consent rates

Screening of individuals took place in September 2022. A total of 240 individuals were screened, the majority were sampled from the governorate of Amman (n=160; 67%), and the remainder from the governorate of Karak (n=80). Primarily Jordanian participants were screened (n=96), with a minority of Syrians (n=64). Of the 240 individuals screened, a total of 83 participants were excluded for reasons including screening negatively on the K10 measure (n=62), reporting imminent suicide (n=10), and could not be contacted (n=11) due to moving away, family refusing participation or declining consent prior to baseline assessment. A further 12 participants refused participation prior to randomization. As a result, 145 were randomized, where 73 were allocated to stepped care and 72 to the single intervention arm (see Figure 1 for exclusion reasons). There were 126 females, and 19 males approximately evenly distributed across intervention arms.

**Fig 1.**
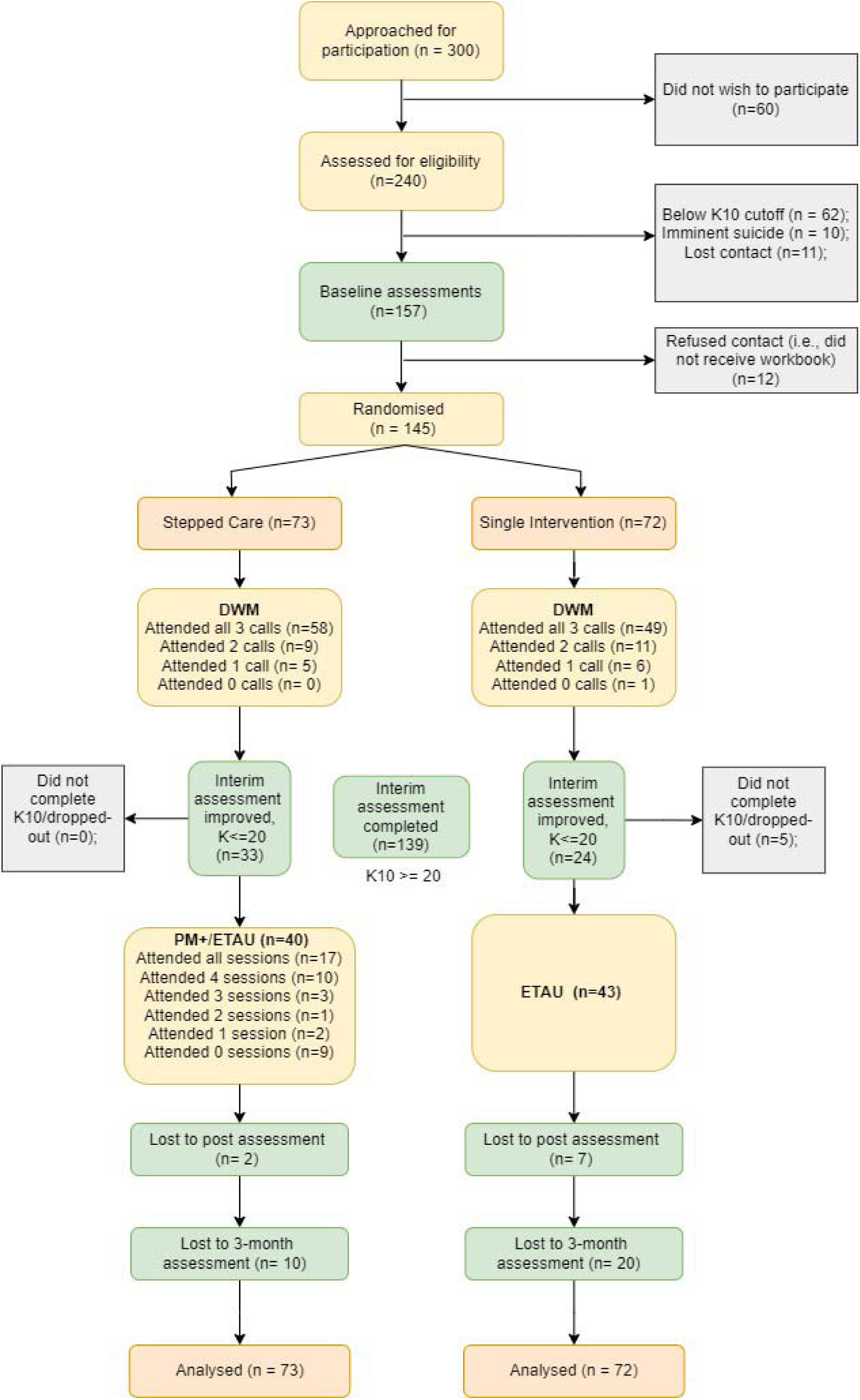
Study flow chart

#### Assessment and retention

Assessments took place between October 2022 and February 2023. Sixty-three (86%) of the 73 participants allocated to stepped care, and 52 (69%) of the 72 participants allocated single intervention, were retained from baseline to three-month follow-up. One hundred and seven (74%) completed all DWM support calls. Seventeen (43%) of the 40 participants allocated to gPM+ completed all five sessions, and 27 (68%) completed at least four gPM+ sessions. Whereas nine (23%) did not attend any of the sessions. At the three-month follow-up time point, data were obtained from 115 participants, with 73% completing all assessments.

#### Fidelity assessments

Fidelity was operationalized as the mean fidelity checklist for the DWM and gPM+ programs. These checklists were used to assess whether key activities were implemented and the competency with which they were completed. For the DWM intervention, all DWM helpers (n=6) agreed to have their support calls audio recorded, and a clinical supervisor scored a random selection of 10 support calls. Fidelity checks indicated that DWM helpers adhered to a 100% of the DWM protocol. For the gPM+ program, a clinical supervisor attended at least two of the five gPM+ sessions to rate the skills of the all gPM+ facilitators (n = 6). These checks indicated that gPM+ facilitators adhered to 92% of the gPM+ protocol.

### B) Acceptability and feasibility

Semi-structured interviews were conducted with assessors (n=6), participants who screened negative on the K10 measure (n = 6), participants who completed the DWM (n=9) and gPM+ (n = 6) interventions, those who dropped out of the gPM+ sessions (n = 2), DWM helpers (n=6), and gPM+ facilitators (n=6) and a clinical supervisor (n=1). Interview questions focused on experiences with assessments, DWM, and gPM+ interventions, and any barriers and enablers to participation in the study. These qualitative interview results have been detailed in Table S3.

#### Screening and Assessments

Assessors generally spoke positively about readily building rapport and trust with intended participants (or beneficiaries), where selection of participants who were existing beneficiaries of IFH greatly facilitated this (Q1.2-1.4). Some participants had changed their contact numbers or residence, and assessors noted that this made it challenging to contact them for baseline assessments following previous screening (Q1.1). Assessors reported that the assessments were perceived as lengthy and arduous by some participants (Q1.5). It was reported that two participants perceived the questions relating to exposure to war (i.e., LEC) as irrelevant and sometimes unacceptable to local Jordanian participants (Q1.6).

#### Experiences with DWM

Overall, participants spoke positively about the strategies in the workbook (Q2.1-2.3) and its therapeutic impact on managing stress and worry and improving their interpersonal relationships (Q2.4). Participants spoke about sharing the workbook with immediate family members, and sometimes engaging in the exercises with them (Q2.4-2.5). Some participants who were parents spoke about reading the workbook together with their adolescent children, spouses, and immediate family members (Q2.2, 2.4). Illiterate participants found the WhatsApp audio messages to be appropriate and useful in facilitating their learning of exercises, where some reported repeatedly revisiting these for the duration of the program (Q2.6-2.7).

Participants spoke positively of the DWM helpers insofar as reporting that they looked forward to receiving support calls within the 5-week period of the program; here, helpers were described as gentle, friendly, and offered a space to talk freely (Q2.8-2.1.0). They reported the support calls were a helpful reminder of learned exercises and how to implement them in their routine lives (Q2.1.1). Some participants wished for calls to be longer in duration (i.e., more than 15 minutes), and all participants expressed a keen interest in having further group support (Q2.1.2-2.1.3). A challenge to engagement was the sense that reading the DWM workbook was a ‘duty’ to some participants (Q2.1.5). Those who dropped out of DWM reported having difficulties with committing to attending support calls due to personal stressors (Q2.1.4). One participant expressed improving upon the program by including homework between calls to aid their preparation (Q2.1.6).

#### Experiences with gPM+

Participants who completed gPM*+* spoke positively about the intervention strategies (Q3.1) and felt skills could be readily implemented in their routine lives (Q3.2), where some reported feeling like they were doing something for themselves (Q3.3). Motivation to attend sessions was facilitated by the need to escape from problems in the home environment, reduce ‘pressure’ felt within themselves, and the general eagerness to learn ways to cope (Q3.5-3.7). Some noted sharing their participation in the sessions with family and friends where this was met with encouragement (Q3.3-3.4). One participant noted that their participation was met with sarcasm from their immediate family members. Participants reported having good rapport with facilitators (Q3.8). One participant reported difficulty in openly speaking about their problem with an unfamiliar person (i.e., gPM+ facilitator) (Q3.9).

Those who dropped out of the gPM+ program (i.e., attended less than three sessions) also spoke positively about the strategies they were exposed to in the sessions attended but cited a varied number of reasons for declining participation. This included moving away or living farther away from the center where gPM+ sessions were held, not having childcare, and having experienced a death in the family (Q3.1.0-3.1.3) during the study. One participant reported wanting individual sessions insofar as needing more attention from the facilitator to address their presenting concerns.

#### Experiences of DWM helpers and gPM+ facilitators

DWM helpers reported that participants found it encouraging to receive support from an unknown other who represented IFH (Q4.1), where this reportedly increased participant adherence (Q4.2). They reported that male participants were relatively less engaged due to circumstantial challenges owing to the olive-harvesting season (Q4.4). The use of WhatsApp voice messages to supplement support calls was reportedly useful in aiding learning by some participants who were illiterate (Q4.5, 4.7-4.8). Some participants requested support calls outside of daytime working hours to accommodate their employment (Q4.3), where this placed undue pressure on some helpers. Helpers reported finding it challenging to deliver workbooks on their own to households comprising of only male participants, where this was managed by having their home visits accompanied by another IFH representative (Q4.6).

gPM*+* facilitators noticed that some participants had difficulty sharing feelings in a group format (Q4.1.2 - 4.1.3). One facilitator stated that having participants of a similar age and diverse backgrounds was generally found to be useful in encouraging learning about one another (Q4.9), but also a cause for delayed rapport building amongst participants. For example, one facilitator noted some participants found it hard at first to openly talk but this was noticed to generally improve by the third session (Q4.1.2). One facilitator stated that participants may have been familiar with other members in the group, where this could have deterred comfort in openly speaking about their problems (Q4.1.4). Facilitators noted that participants who were employed found it difficult to attend sessions regularly (Q4.1.5). For others, reimbursement for transportations costs may have been insufficient (e.g., those residing in Karak and travelling to the IFH center), and this may have reduced their adherence (Q4.1.6).

#### Experiences in clinical supervision

Some challenges were reported during supervision sessions. First, DWM helpers were observed to experience challenges with readily responding to participants who expressed religious or spiritual coping (e.g., prayers) insofar as guiding participants on how to use DWM strategies. To this end, it was observed that gPM+ facilitators were relatively more experienced in responding to such ways of coping and reported that this required further attention during training for the fully powered RCT. Additionally, helpers reported receiving many queries related to wanting financial support, and to this end, requested more training on how to respond to such problems during DWM support calls. Overall, gPM+ and DWM non-specialists were characterized as motivated to deliver the interventions to participants. Some gPM+ facilitators declined to attend regular clinical supervision due to competing work demands at IFH, and to this end, were reported to miss out on supervision opportunities relative to others without such work demands.

### C) Estimated clinical outcomes

There were no significant differences in socio-demographic characteristics (Table 1) and exposure to lifetime traumatic events (Table 2) across intervention arms. Nearly a third of participants had been exposed serious accidents, life threatening illnesses, serious physical injury, been in danger during flight from warzones, forced separation from family members, and experienced a lack of food or water. Table 3 presents linear mixed model analyses for primary outcomes including depression and anxiety (HSCL-25), and secondary outcomes including wellbeing (WHO-5), agency (State Hope Scale), functional impairment (WHODAS v2.0), quality of life (EURO-QoL), and self-identified problems (PSYCHLOPS). Table S1 presents linear mixed model analyses for all outcomes for those who were retained at the 3-month follow-up only (see supplement). Table S2 presents the frequency of self-identified problems reported as primary (PSYCHLOPS Q1a) and other (PSYCHLOPS Q2a) problems. No serious adverse events were reported during the trial.

**Table 1.**
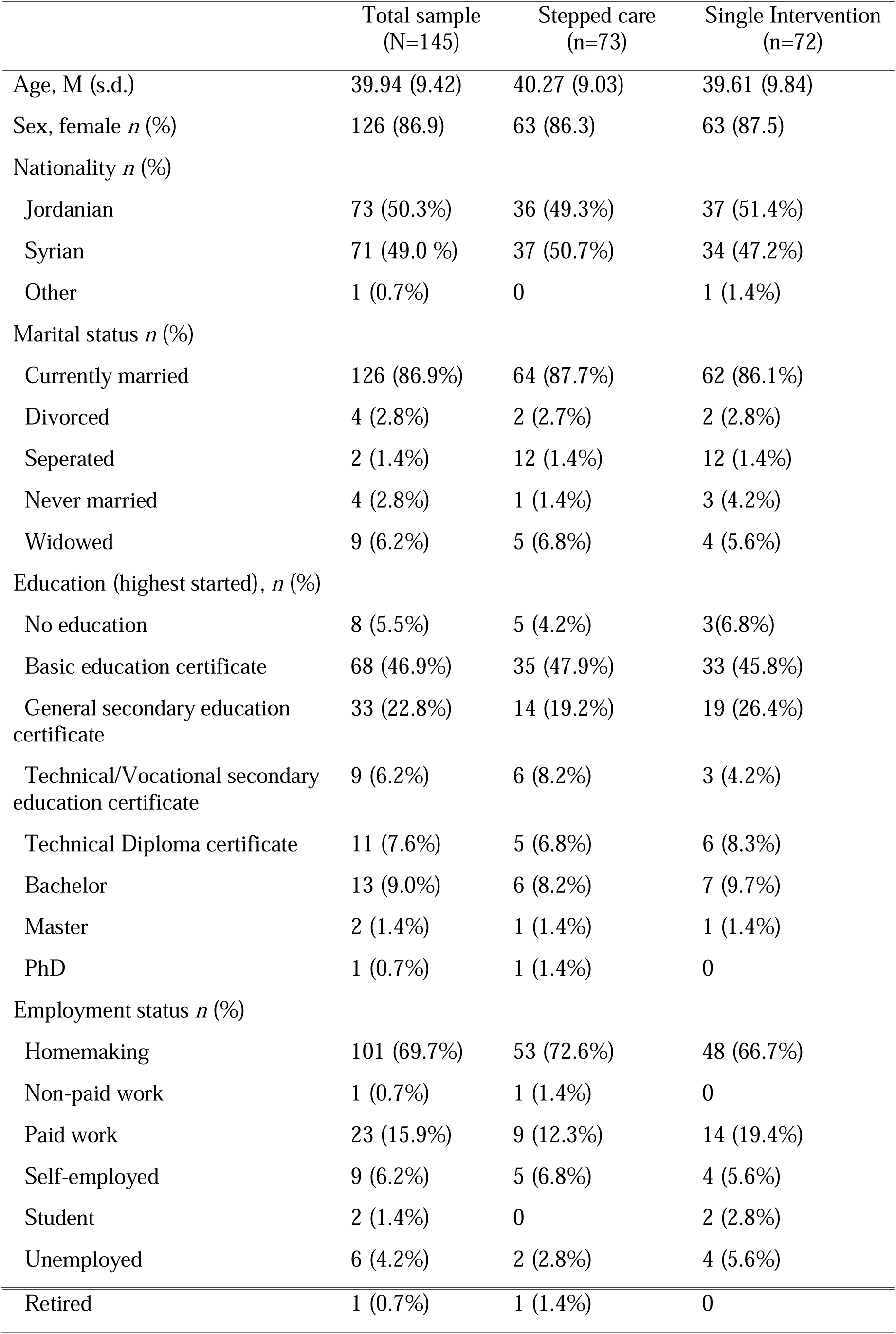
Baseline demographic characteristics.

**Table 2.**
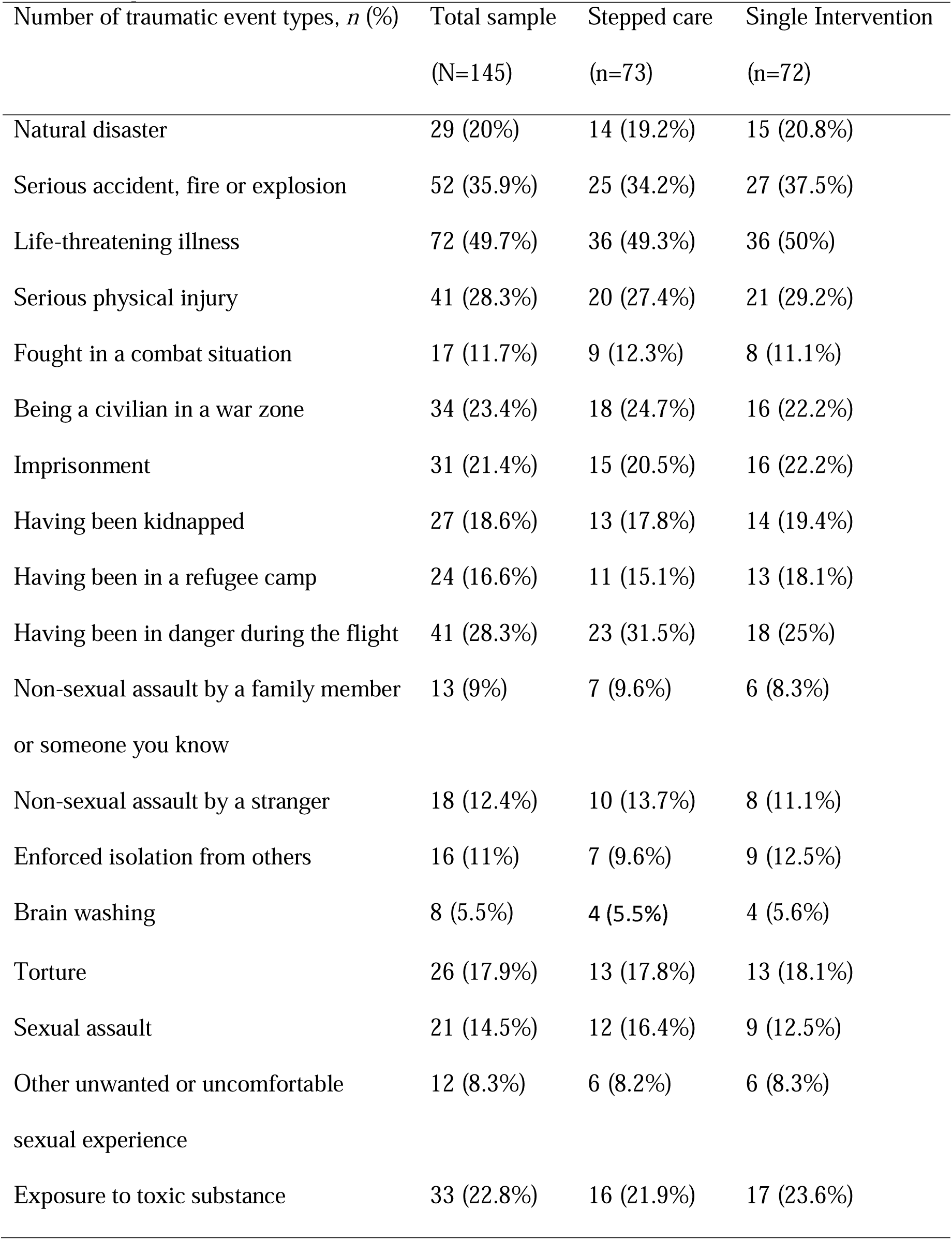
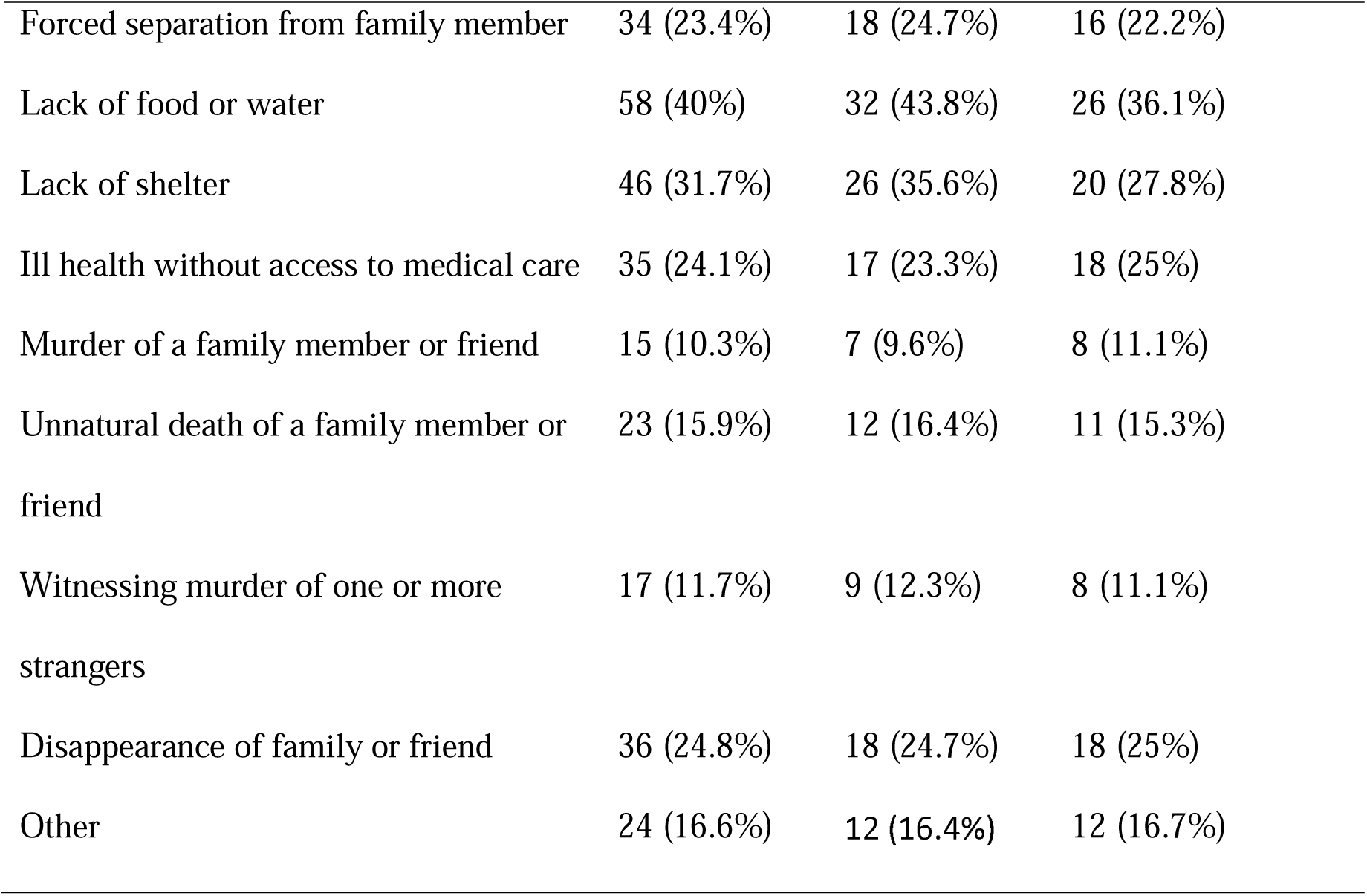
Exposure to traumatic events.

**Table 3.**
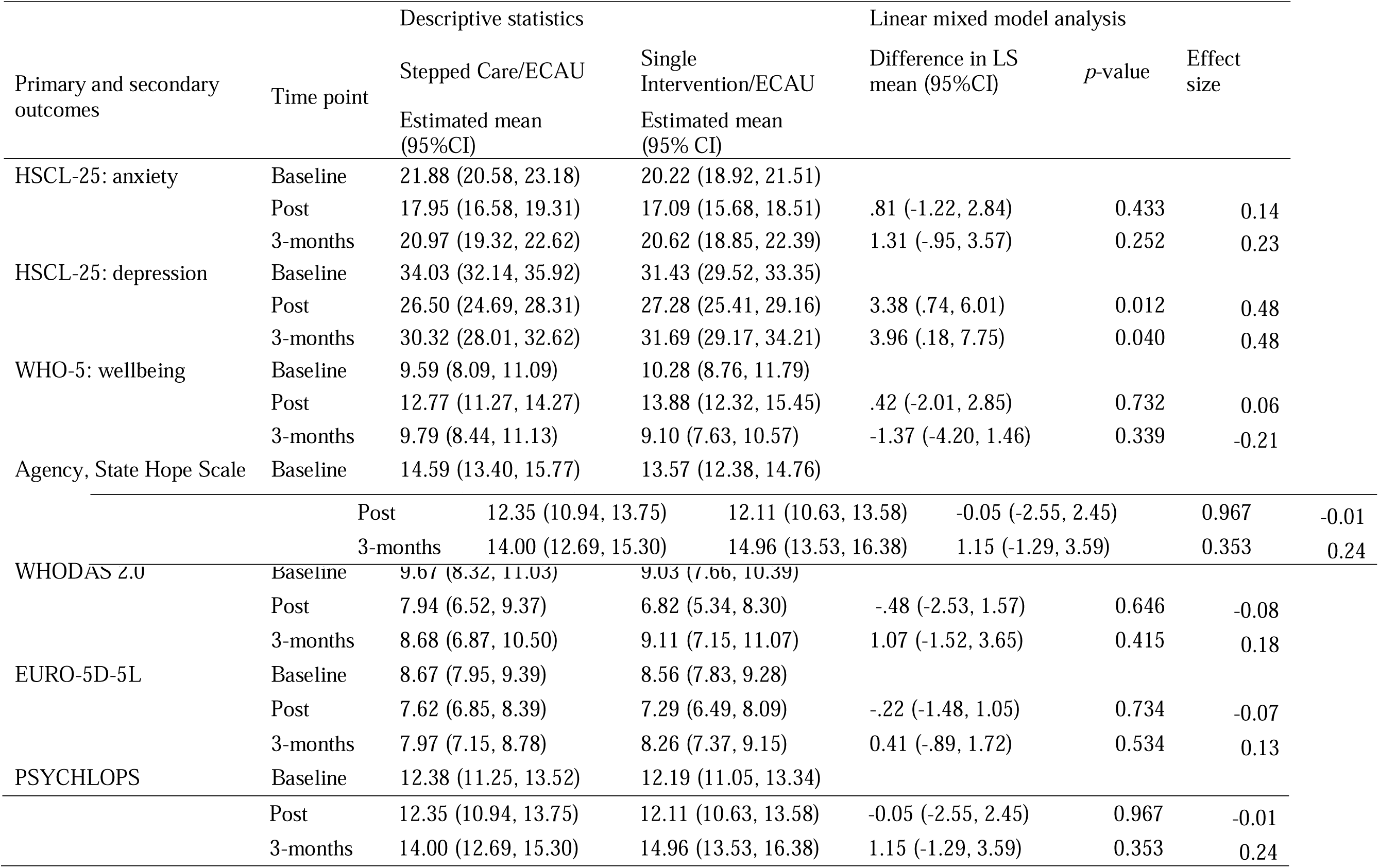
Summary of clinical outcomes.

Although depression (*F*(2, 126) = 39.50, *p*<0.001) and anxiety (*F*(2, 123) = 27.55, *p*<0.001) significantly reduced over time, there were no overall significant differences between baseline and follow-up (*t*(122)=1.81, *p* = 0.073). Reductions in depression differed by group and time, where the stepped care arm experienced greater reductions in symptoms from baseline to post (*mean difference* 3.38, [*95% CI*, .74, 6.02, *p* = 0.012]), and to follow-up (*mean difference* 3.96, [*95% CI*, .18, 7.75], *p*=0.04) relative to the single intervention arm. Similar between group changes were not found for anxiety from baseline to post (*mean difference* .81, [*95% CI*, 1.22, 2.84], *p*>0.05), and to follow-up (*mean difference* 1.31, [*95% CI*, -.95, 3.57], *p*>0.05). There were no other between group changes on all secondary outcome measures.

## Discussion

The aim of the current pilot study was to assess the feasibility and acceptability of a stepped care approach to delivering brief psychological interventions to adults affected by adversity in Jordan. Participants comprising of both Jordanians and Syrians had been exposed to a range of traumatic events at the time of the trial. Overall, findings suggest that the DWM and gPM+ interventions were found to be acceptable, beneficial for participants, and feasible to deliver within the Jordanian context. To this end, financial problems, emotional problems, and interpersonal problems, respectively were the most highly rated across intervention arms. Nearly 80% of participants were retained in the study from baseline assessment to the three-month follow-up assessment. Adherence in the DWM intervention was high with over 83% and 92% of participants attending at least two support calls in the single-intervention and stepped care arms, respectively. Adherence in the gPM*+* intervention was relatively lower with 68% of participants attending at least four sessions. There were no serious adverse events that took place during the trial suggesting that this model of care is safe for delivery within the Jordan context.

As the first study in a LMIC setting to assess the delivery of a brief guided self-help intervention, followed by a more intensive group program, the current study findings suggest that the DWM and gPM+ interventions may be beneficial in teaching participants ways to cope with their distress and that it provided an increased understanding of how to manage routine problems in their lives. Importantly, participants expressed a need for these interventions, where delivery of these by non-specialists was perceived to be acceptable and appropriate insofar assisting them with their practice of learned strategies and increasing their knowledge gain from the interventions. Across both interventions, participants reported an increase in their interpersonal interactions including open discussions with immediate family members, friends, and families about their participation in the interventions. To this end, some participants found practicing strategies with others to be helpful where this largely involved encouragement from close others. The group format of gPM+ received mixed feedback as reported by the gPM+ facilitators where some participants were reported to experience difficulties with building rapport readily, whereas others did not experience this challenge. Reasons for non-adherence included been engaged in employment, difficulties with arranging childcare, and sudden illnesses that together precluded attendance. To this end, the recruitment of males into the study was a notable challenge because these participants were often engaged in active employment or were otherwise occupied with the olive-harvesting season that limited their full participation. The overall feasibility and acceptability of conducting the stepped care program with males suggests that whilst possible to conduct, these barriers to participation may need to be addressed to improve upon current adherence rates. Recommendations from non-specialists and participants (for a definitive trial) to address barriers and improve adherence to interventions included accommodations for childcare for the duration gPM+ sessions, increasing reimbursements for those travelling from a distance to attend gPM+ sessions, offering alternate group allocations for those in active employment, and providing gPM+ facilitators with IFH with opportunities to attend regular supervision. To this end, it was recommended by program staff for gPM+ facilitators to be allotted time to attend supervision for the duration of the trial. A strength of this study was the use of existing IFH beneficiary lists to recruit participants where this was found to both increase acceptability and continued adherence with the interventions. This view was endorsed by program staff, non-specialists, and participants.

Overall, both intervention arms appeared to benefit from DWM and gPM+ interventions over time, although we note the design did not include a no-treatment comparator. The stepped care arm greatly benefitted from a reduction in depressive symptoms relative to those only receiving a single intervention. This finding must be interpreted with caution as the study was not adequately powered to detect significant results. At the same time, qualitative analyses support this finding in revealing that most participants experienced subjective improvements after participation in the DWM and gPM*+* interventions. If replicated in a fully powered trial, this finding would provide initial support for a stepped model of care for allocation of limited resources in a LMIC setting.

Our study had several limitations. First, participants were recruited from existing IFH beneficiary lists who were help-seeking and potentially more open to receiving psychological support because of this affiliation. The findings are likely to be different for those individuals not acquainted with a known organization such as IFH insofar as a potentially higher need to build trust and rapport prior to engaging in recruitment. Second, drop-out within the single intervention arm was twice the number of those that dropped out in the stepped care arm, and this difference may have influenced findings as seen the lack of significant finding in depression symptoms when we conduced completer analyses (see Table S1). At the same time, it is acknowledged that the trial was not sufficiently powered to detect meaningful differences. Third, some gPM+ facilitators did not attend supervision regularly due to competing role demands within the organization. Whilst this appeared to minimally impact competency and fidelity to the gPM+ intervention, dedicated time to commit to the duration of the gPM+ intervention is recommended for the fully powered trial.

In conclusion, lessons learned from this feasibility trial suggests that there was sufficient interest and need for brief psychological interventions including DWM and gPM+ within a stepped care framework in Jordan. Whilst these programs can be competently delivered by non-specialists, receiving regular supervision is crucial. Taken together, these results suggest that a stepped model of psychological care as delivered by non-specialists is feasible, well-accepted and a safe option for adults affected by adversity in Jordan. A fully powered trial is currently underway and will seek to address barriers to adherence noted above and further establish the clinical and cost-effectiveness of this framework for the Jordan context.

## Supporting information

CONSORT checklist

## Data Availability

Data will be available upon request.

## Acknowledgments

We are grateful to the dedicated team at the Institute for Family Health (IFH), the study participants, facilitators, clinical supervisors, and professionals who provided their time and input into the formative research guiding this trial.

## Financial support

This study was supported by a r2hc (research for health in humanitarian crises) elrha grant. Grant number #47455. The funder had no role in study design, data collection and analysis, decision to publish or preparation of the manuscript.

## Conflict of Interests

None.

## Ethical standards

This trial received ethical approval through the University of Jordan School of Nursing Research Ethics Committee (number: PF.22.10). The authors assert that all procedures contributing to this work comply with the ethical standards of the relevant institutional committee (as per the University of Jordan School of Nursing Research Ethics Committee regulations).

## Availability of data and materials

Data will be available upon request.

## Notes

### Competing Interest Statement

The authors have declared no competing interest.

### Clinical Trial

ACTRN12621000189820p

## References

1. Epping-Jordan JE, Harris R, Brown FL, et al. Self-Help Plus (SH+): a new WHO stress management package. World Psychiatry 2016;15(3):295–96. doi: 10.1002/wps.20355

2. Purgato M, Carswell K, Tedeschi F, et al. Effectiveness of Self-Help Plus in Preventing Mental Disorders in Refugees and Asylum Seekers in Western Europe: A Multinational Randomized Controlled Trial. Psychother Psychosom 2021;90(6):403–14. doi: 10.1159/000517504 [published Online First: 20210720]

3. Acarturk C, Uygun E, Ilkkursun Z, et al. Effectiveness of a WHO self-help psychological intervention for preventing mental disorders among Syrian refugees in Turkey: a randomized controlled trial. World Psychiatry 2022;21(1):88–95. doi: 10.1002/wps.20939 [published Online First: 2022/01/12]

4. Purgato M, Turrini G, Tedeschi F, et al. Effectiveness of a stepped-care programme of WHO psychological interventions in migrant populations resettled in Italy: Study protocol for the RESPOND randomized controlled trial. Frontiers in Public Health 2023;11 doi: 10.3389/fpubh.2023.1100546

5. Dawson KS, Bryant RA, Harper M, et al. Problem Management Plus (PM+): a WHO transdiagnostic psychological intervention for common mental health problems. World psychiatry : official journal of the World Psychiatric Association (WPA) 2015;14(3):354–57. doi: 10.1002/wps.20255

6. Bryant RA, Schafer A, Dawson KS, et al. Effectiveness of a brief behavioural intervention on psychological distress among women with a history of gender-based violence in urban Kenya: A randomised clinical trial. PLoS Med 2017;14(8):e1002371. doi: 10.1371/journal.pmed.1002371 [published Online First: 20170815]

7. Rahman A, Hamdani SU, Awan NR, et al. Effect of a multicomponent behavioral intervention in adults impaired by psychological distress in a conflict-affected area of Pakistan: A randomized clinical trial. JAMA - Journal of the American Medical Association 2016;316(24):2609–17. doi: 10.1001/jama.2016.17165

8. Bryant RA, Bawaneh A, Awwad M, et al. Effectiveness of a brief group behavioral intervention for common mental disorders in Syrian refugees in Jordan: A randomized controlled trial. PLoS Med 2022;19(3):e1003949. doi: 10.1371/journal.pmed.1003949 [published Online First: 20220317]

9. Jordans MJD, Kohrt BA, Sangraula M, et al. Effectiveness of Group Problem Management Plus, a brief psychological intervention for adults affected by humanitarian disasters in Nepal: A cluster randomized controlled trial. PLoS medicine 2021; 18(6). http://europepmc.org/abstract/MED/34138875https://journals.plos.org/plosmedicine/article/file?id=10.1371/journal.pmed.1003621&type=printablehttps://doi.org/10.1371/journal.pmed.1003621https://europepmc.org/articles/PMC8211182https://europepmc.org/articles/PMC8211182?pdf=render (accessed 2021/06//).

10. Rahman A, Khan MN, Hamdani SU, et al. Effectiveness of a brief group psychological intervention for women in a post-conflict setting in Pakistan: a single-blind, cluster, randomised controlled trial. The Lancet 2019;393(10182):1733–44. doi: 10.1016/S0140-6736(18)32343-2

11. Schäfer SK, Thomas LM, Lindner S, et al. World Health Organization’s low-intensity psychosocial interventions: a systematic review and meta-analysis of the effects of Problem Management Plus and Step-by-Step. World Psychiatry 2023;22(3):449–62. doi: 10.1002/wps.21129 [published Online First: 2023/09/15]

12. Salomonsson S, Santoft F, Lindsäter E, et al. Stepped care in primary care - guided self-help and face-to-face cognitive behavioural therapy for common mental disorders: a randomized controlled trial. Psychological medicine 2018;48(10):1644–54. doi: 10.1017/s0033291717003129

13. Khatib HE, Alyafei A, Shaikh M. Understanding experiences of mental health help-seeking in Arab populations around the world: a systematic review and narrative synthesis. BMC Psychiatry 2023;23(1):324. doi: 10.1186/s12888-023-04827-4 [published Online First: 20230509]

14. Keyan D, Habashneh R, Akhtar A, et al. Evaluating a Stepped Care Model for Psychological Support for Adults Affected by Adversity: Study Protocol for a Randomised Controlled Trial in Jordan. BMJ Open 2024;14(2):e078091. doi: 10.1136/bmjopen-2023-078091 [published Online First: 2024/02/28]

15. Kessler RC, Barker PR, Colpe LJ, et al. Screening for serious mental illness in the general population. Arch Gen Psychiatry 2003;60(2):184–9. doi: 10.1001/archpsyc.60.2.184

16. WHO. Doing What Matters in Times of Stress: Geneva: World Health Organziation, 2021.

17. Applied Mental Health Research Group. Design, implementation, monitoring, and evaluation of cross-cultural HIV-related mental health and psychosocial assistance programs: a user’s manual for researchers and program implementers (adult version): Baltimore: Johns Hopkins University;, 2013.

18. Akhtar A, Engels M, Bawaneh A, et al. Cultural Adaptation of a Low-Intensity Group Psychological Intervention for Syrian Refugees. Intervention 2021;19(1):48–57. doi: 10.4103/intv.Intv_38_20

19. Derogatis LR, Lipman RS, Rickels K, et al. The Hopkins Symptom Checklist (HSCL): a self-report symptom inventory. Behav Sci 1974;19(1):1–15. doi: 10.1002/bs.3830190102

20. Fares S, Dirani J, Darwish H. Arabic validation of the hopkins symptom checklist-25 (HSCL) in a Lebanese sample of adults and older adults. Current Psychology 2021;40(6):2980–87. doi: 10.1007/s12144-019-00240-x

21. Krieger T, Zimmermann J, Huffziger S, et al. Measuring depression with a well-being index: Further evidence for the validity of the WHO Well-Being Index (WHO-5) as a measure of the severity of depression. Journal of Affective Disorders 2014;156:240–44. doi: 10.1016/j.jad.2013.12.015

22. Ustun TB, Kostanjsek, N., Chatterji, S., Rehm, J editor. Measuring health and disability : manual for WHO Disability Assessment Schedule (WHODAS 2.0). World Health Organization., 2010.

23. Weathers FW, Blake DD, Schnurr PP, et al. The Life Events Checklist for DSM-5 (LEC-5) 2013 [Available from: Instrument available from the National Center for PTSD at www.ptsd.va.gov.

24. Herdman M, Gudex C, Lloyd A, et al. Development and preliminary testing of the new five-level version of EQ-5D (EQ-5D-5L). Qual Life Res 2011;20(10):1727–36. doi: 10.1007/s11136-011-9903-x [published Online First: 20110409]

25. Ashworth M, Shepherd M, Christey J, et al. A client-generated psychometric instrument: The development of ‘PSYCHLOPS’. Counselling and Psychotherapy Research 2004;4(2):27–31. doi: 10.1080/14733140412331383913

26. Snyder CR, Sympson SC, Ybasco FC, et al. Development and validation of the State Hope Scale. J Pers Soc Psychol 1996;70(2):321–35. doi: 10.1037//0022-3514.70.2.321

27. Schleider JL, Mullarkey MC, Fox KR, et al. A randomized trial of online single-session interventions for adolescent depression during COVID-19. Nature Human Behaviour 2022;6(2):258–68. doi: 10.1038/s41562-021-01235-0

28. NVivo (Version 14) [program], 2023.

