## Supplement for "A stepped-care programme of brief psychological interventions for adults affected by adversity in Jordan: Lessons from a pilot randomised controlled trial in Jordan"

**Table S1.** Summary of clinical outcomes for those completing the 3-month follow-up

|  |  | Descriptive statistics | | Linear mixed model analysis | | |
| --- | --- | --- | --- | --- | --- | --- |
| Primary and secondary outcomes | Time point | Stepped Care (DWM + gPM+/ ETAU) | Single Intervention (DWM+ETAU) | Difference in LS mean (95%CI) | *p*-value | Effect size |
|  |  | Estimated mean (95%CI) | Estimated mean (95% CI) |  |  |  |
| HSCL-25: anxiety | Baseline | 21.81 (20.48, 23.13) | 21.00 (19.56, 22.45) |  |  |  |
|  | Post | 17.52 (16.19, 18.86) | 17.12 (15.64, 18.59) | .39 (-1.74, 2.53) | 0.720 | 0.07 |
|  | 3-months | 20.76 (19.12, 22.40) | 20.96 (19.16, 22.77) | 1.00 (-1.29, 3.29) | 0.387 | 0.19 |
| HSCL-25: depression | Baseline | 34.04 (32.08, 36.00) | 32.63 (30.47, 34.80) |  |  |  |
|  | Post | 26.49 (24.57, 28.42) | 27.83 (25.71, 29.94) | 2.74 (-.19, 5.68) | 0.067 | 0.35 |
|  | 3-months | 30.32 (28.00, 32.63) | 32.06 (29.51, 34.60) | 3.15 (-.72, 7.02) | 0.110 | 0.40 |
| WHO-5: wellbeing | Baseline | 9.56 (8.04, 11.07) | 8.81 (7.14, 10.47) |  |  |  |
|  | Post | 12.62 (11.02, 14.22) | 13.92 (12.16, 15.68) | 2.05 (-.55, 4.66) | 0.122 | 0.34 |
|  | 3-months | 9.75 (8.40, 11.10) | 9.17 (7.69, 10.66) | .17 (-2.73, 3.08) | 0.905 | 0.03 |
| Agency, State Hope Scale | Baseline | 14.70 (13.46, 15.94) | 13.13 (11.77, 14.50) |  |  |  |
|  | Post | 16.54 (15.37, 17.71) | 16.65 (15.37, 17.94) | 1.68 (-.47, 3.83) | 0.125 | 0.34 |
|  | 3-months | 13.95 (12.61, 15.29) | 14.06 (12.58, 15.53 | 1.67 (-.81, 4.15) | 0.186 | 0.33 |
| WHODAS 2.0 | Baseline | 9.75 (8.32, 11.17) | 9.90 (8.33, 11.48) |  |  |  |
|  | Post | 7.87 (6.37, 9.38) | 6.88 (5.23, 8.54) | -1.15 (-3.37, 1.08) | 0.309 | -0.20 |
|  | 3-months | 8.68 (6.84, 10.52) | 9.44 (7.42, 11.47) | .60 (-2.03, 3.23) | 0.651 | 0.11 |
| EURO-5D-5L | Baseline | 8.63 (7.87, 9.39) | 9.02 (8.18, 9.86) |  |  |  |
|  | Post | 7.71 (6.92, 8.51) | 7.21 (6.33, 8.09) | -.89 (-2.21 .44) | 0.188 | -0.29 |
|  | 3-months | 7.97 (7.14, 8.79) | 8.40 (7.50, 9.31) | .05 (-1.29, 1.39) | 0.940 | 0.02 |
| PSYCHLOPS | Baseline | 12.49 (11.35, 13.64) | 12.77 (11.51, 14.03) |  |  |  |
|  | Post | 12.81 (11.28, 14.34) | 12.04 (10.36, 13.72) | (-1.05) (-3.89, 1.79) | 0.466 | -0.23 |
|  | 3-months | 14.14 (12.83, 15.46) | 14.96 (13.51, 16.41) | .54 (-2.00, 3.08) | 0.674 | 0.12 |

**Table S2.** Frequency of self-identified primary (PSYCHLOPS 1a) and other (PSYCHLOPS 2a) problems

|  | Stepped Care | | Single Intervention | |
| --- | --- | --- | --- | --- |
| Problem code | Primary problem, *N* (%) | Other Problem, *N* (%) | Primary problem, *N* (%) | Other Problem, *N* (%) |
| Financial problems | 34 (46.6%) | 15 (20.5%) | 26 (36.1%) | 12 (16.7%) |
| Psychological/emotional | 17 (23.3%) | 6 (8.2%) | 20 (27.8%) | 4 (5.6%) |
| Interpersonal problems | 9 (12.3%) | 2 (2.7%) | 12 (16.7%) | 5 (6.9%) |
| Poor health/Health problems | 1 (1.4%) | 8 (11.0%) | 2 (2.8%) | 6 (8.3%) |
| Other's health problems | 4 (5.5%) | 5 (6.8%) | 5 (6.9%) | 3 (4.2%) |
| Unemployment | 4 (5.5%) | 2 (2.7%) | 2 (2.8%) | - |
| Other | 1 (1.4%) | 2 (2.7%) | 1 (1.4%) | 2 (2.8%) |
| None (no problem coded) | 3 (4.1%) | 32 (43.8%) | 3 (4.2%) | 40 (55.6%) |

**Table S3.** Qualitative feedback

| Domain: | Theme | # | Quote |
| --- | --- | --- | --- |
| Experiences with Assesments | Trial procedures | Q1.1 | “The issue in Sweileh was that we had to choose individuals from the same area. Still, we were calling different individuals from the available data and finding out that, they moved from the registered address to another one.” |
|  | Rapport with beneficiaries | Q1.2  Q1.3  Q1.4 | “There was a kind of fast acceptance from people due to the Foundation’s reputation.”;  “Trust developed, and sometimes we would pause and listen to their issues, and the woman spoke comfortably”;  “ I called this phase, the trust phase, At this level you have to build trust between you and the beneficiaries and talk more about the project, .. I think this is the most important phase, it was not just asking a question and waiting for answers in boring way, no it was interactive discussions.” |
|  | Length of questionnaires | Q1.5 | “The questionnaire's length, the repetition of certain questions, and the need for certain questions.” |
|  | Appropriateness of questions | Q1.6 | “The questions must be more flexible, as both Syrian and Jordanian are in this project, but each one lives in a different society and faces different challenges, and … there are general question that should be specified to the Syrians only, about the war or if you were kidnapped” |
| Experiences with DWM | Perceived benefit | Q2.1  Q2.2  Q2.3 | “ I stopped exaggerating things; for example, when my daughter came late from school, I used to have negative thoughts that she might be kidnapped or lost; my thoughts became better and I began thinking positively”;  “ The book does not remove the problem rather, it teaches you how to accept it and live with it, and that what you have now is a grace. I still until now apply the exercises; they are really like a lifestyle to me and my son”;  “ I started setting aside time for myself, telling myself that life isn’t just work, work, work. I would sit and relax with a cup of coffee. After lunch, I would make a cup of coffee and sit with my husband” |
|  | Sharing with family | Q2.4  Q2.5 | “ My husband, my young daughters, and I do the exercises together. When I do a movement, they imitate me. My second-grade daughter tells me, "Let’s read the book," and she sits and reads it to me. Sometimes, while we’re having coffee, my husband asks me to sit and tell him how I relax and do the exercises”;  “I told my sisters, and they saw the book. One of them read it and wanted to participate by reading the book as she loved it” |
|  | Support for participants who could not read | Q2.6  Q2.7 | “ when I open the recordings, my wife listens to them, she does not know how to read so she listens with me”  “Frankly, my situation prevented me from reading the book, but I benefited from the audio recordings I used to open and pause, all of them were great” |
|  | Rapport with helper and support calls | Q2.8  Q2.9  Q2.1.0  Q2.1.1 | “ The idea is that somebody is listening to you ..someone that does not judge, someone that does not focus on your mistakes, does not want to have any negative thoughts about you”;  “The girls are educated and master their work. They have the ability to know all the psyches of people. I have no idea what their domain is but they have that ability to know your situation.”;  “ It is that they explained what we do not understand, we benefit from it, and we feel comfortable. I mean, we feel very comfortable, and sometimes we forget things and they remind us of them”  “Getting my attention to things I didn’t notice before, as I reduced the number of my daily cigarettes. The earthquake issue affected us so much, the book helped me in how to deal with shocks.” |
|  | Requests for further support | Q2.1.2  Q2.1.3 | “ We need more support; we want group sessions.”  “ It would be good to have a call every week.” |
|  | Challenges to participation | Q2.1.4  Q2.1.5 | “because of my mother’s illness I cannot attend every day as I go to the hospital and I have my children to take care of them and I have to stay home sometimes”;  “my problem revolves around the book, for example: I do not want to feel that I have a continuous assignment, and when I do not read I feel embarrassed not knowing what to say. I sometimes have issues. the girl used to say "No problem" and she gave me time and explained to me, but I used to feel responsible and I had an assignment to do.” |
|  | Suggestions for improvement | Q2.1.6 | “It would be great if you could send us topics like homework so we can prepare for the session”; |
| Experiences with gPM+ | Perceived benefit | Q3.1  Q3.2 | “ I learned from the sessions that the smallest problem shall not have quick reaction, I now think about it first and decide whether it’s big or huge. I used to consider all problems bigger than they are before,”;  “[daughter] I shared with her the method for solving her problem. She told me that her friend is not talking to her, I asked her about the reason behind that and whether she made her upset, and I told her that she needs to know why and gave her some advices to recognize the problem and solve it” |
|  | Family engagemnet | Q3.3  Q3.4 | “ my daughters told me to attend the session and help myself”;  “ I shared it with my friends to reduce the size of problems”; |
|  | Reasons for engaging | Q3.5  Q3.6  Q3.7 | “ escaping the house problems and burdens”;  “ exchange points of view”;  “ we benefit from the sessions and learn new things” |
|  | Rapport with facilitators | Q3.8 | “ she was kind, optimistic, and cooperative, and the things we didn't understand, she used to repeat it” |
|  | Experiences with the group format | Q3.9 | “.. in collective session there are things that we cannot talk about in front of others, like secrets”;  “ when I share my secrets with someone I think we won’t refer to each other later” |
|  | Reasons for dropping out | Q3.1.0  Q3.1.1 | “I wanted to continue with you, but I had conditions, and it became difficult for me to reach out. I became sick, then my son got sick, and my residence became far from the center.”;  “when I was residing close to you that was appropriate, but now it is difficult, especially since we are residing very far from the center and the financial condition does not allow me to pay for transportation” |
|  | Challenges | Q3.1.2  Q3.1.3 | [distance from center]: “sometime I do not have money to attend”; “I mean we obtained transportation allowance but not at the time of first nor second sessions”;  [childcare needs]:“if I find a place where I can leave my son” |
| Experiences of DWM Helpers | Engagement | Q4.1  Q4.2 | “The thing that encouraged people to talk to me was that they didn’t know me. To them, I was a stranger.”  “People continued because of the foundation’s name. Some thought the task was difficult but were reassured it was just a book.” |
|  | Challenges with providing support | Q4.3  Q4.4  Q4.5  Q4.6 | “Their working hours did not fit the sessions’ time, so they requested evening sessions. We conducted some calls during the day and some at night, which put pressure on us.”;  “Some did not read and tried to convince us that they did, especially during some seasons like in the olive harvesting time.”  “The negatives were that some could not read due to illiteracy, so I sent voice recordings. Despite this, some benefited while others did not absorb the information.”;  “As girls, we visited some houses during the field visits where there were only men present, or some houses seemed suspicious, so I proposed that it would be preferable to have a young man from the Foundation accompany us” |
|  | Adhoc adaptations | Q4.7  Q4.8 | “For the illiterate people, I discussed the next idea with them to encourage and motivate them, advising them to apply the exercises correctly and praising them when they do so.”;  “I found that sending voice recordings with the exercise idea was helpful, and in terms of time, it seemed appropriate.” |
| Experiences of gPM+ facilitators | Perceived benefits | Q4.9  Q4.1.0  Q4.1.1 | “This mixture of Jordanians and Syrians, in my opinion, is excellent since it allows people to get to know one another better and learn about one another's cultures. It follows that the group got along well.”  “The good thing was that they were able to realize how .. their difficulties were in comparison to others' problems, which gave them hope ..”;  “Some said they wanted to learn new things, while others said they wanted to discover their inner strength and gain new experiences. Still others said they wanted to discuss their problems with other people” |
|  | Group format | Q4.1.2  Q4.1.3 | “In the first two sessions, they were not very comfortable speaking about everything they suffered from, but in the fourth session, they started to share their issues openly.”  “the men group it was different from the women’s group, it was hard to make them speak .., some of them may share his problem while the other may remain silent, but after the fourth session they changed” |
|  | Concerns about privacy | Q4.1.4 | “I initially believed that none of them knew the others, but I later discovered that ten of them were already acquainted. This might potentially hinder their ability to communicate openly, there are friends, neighbors, and daughters-in-law in my group. It's important to recognize the dynamics of our society”. |
|  | Challenges with transportation and attendance | Q4.1.5  Q4.1.6 | “ believe that workers … if their work schedules conflict with the program's schedule. Therefore, it would be preferable to select individuals who are not employed”;  “ in Karak, people ask for more than three dinars because some places are far from the Al-Thiniyah area. As my colleague mentioned, they need to be paid more than they received for the transportation.” |
